# Automating Evaluation of LLM-generated Responses to Patient Questions about Rare Diseases

**DOI:** 10.1101/2025.10.06.25337181

**Authors:** Min Zhao, Inez Y. Oh, Aditi Gupta, Sally Cohen-Cutler, Kathryn M. Harmoney, Albert M. Lai, Bryan A. Sisk

## Abstract

**Objectives:** Patients with rare diseases often struggle to find accurate medical information, and large language model (LLM)-based chatbots may help meet this need. However, evaluating LLM-generated free-text answers typically requires physician review, which is time-consuming and difficult to scale. This study compared traditional natural language processing (NLP) metrics to emerging LLM-based evaluation approaches for assessing answer quality in the context of Complex Lymphatic Anomalies (CLAs).

**Materials and Methods:** We compiled 25 common patients’ questions about CLAs and generated 175 responses to these questions from seven LLMs. Three expert physicians scored these responses for accuracy. We compared these physician-assigned scores with automated scores, generated by four NLP sentence similarity metrics (BLEU, ROUGE, METEOR, BERTScore) and six LLM evaluators (GPT-4, GPT-4o, Qwen3-32B, DeepSeek-R1-14B, Gemma3-27B, LLaMA3.3-70B). We examined both LLM-based scoring with and without reference answers (reference-guided vs. reference-free). We calculated Spearman, Phi, and Kendall’s Tau correlation coefficients to assess alignment between automated and physician-assigned scores.

**Results:** LLM-based evaluation demonstrated stronger alignment with physician-assigned scores than NLP metrics. The reference-guided GPT-4 evaluator achieved the highest correlation with physician-assigned scores (ρ=0.758), followed by GPT-4o (ρ=0.727). NLP metrics showed weak to moderate correlations with physician-assigned scores (ρ=0.240–0.403). Reference-guided scoring outperformed reference-free methods.

**Discussion:** Reference-guided LLM-based evaluation methods approximate expert physicians’ judgment better than traditional NLP metrics, offering an effective, scalable approach for assessing LLM-generated responses to patient questions about rare disease.

**Conclusion:** LLM-based evaluation, particularly reference-guided scoring with GPT models, can support the scalable development and evaluation of LLM-based rare disease-specific chatbot systems.

## INTRODUCTION

Rare diseases, as defined by the U.S. National Institutes of Health (NIH), are conditions that affect fewer than 200,000 people in the United States.[1] Although individually rare, these diseases collectively affect millions of individuals worldwide and impose a substantial burden on patients and their families.[2–4] Rare disease patients experience barriers to accessing expert physicians to manage their care, and often struggle to find accurate and reliable information about their condition.[5–11] In this study, as a model system for understanding information needs and communication barriers in rare diseases, we focused on Complex Lymphatic Anomalies (CLAs), a group of four rare diseases characterized by abnormal lymphatic development that can cause high morbidity and lifelong complications.[12] Prior studies have shown that accessing high-quality information is essential for patients and families affected by CLAs to receive optimal care.[5,6,13] In fact, patients who report receiving better information about their disease also report better mental health, physical health, and ability to navigate the healthcare system.[14,15] Given the dearth of expert clinicians and lack of high-quality information, there is a critical need to develop accurate chatbots that can support the information needs of patients and families affected by CLAs.

Recent advancements in large language models (LLMs) offer a promising opportunity to develop scalable, patient-centered AI chatbot systems tailored to rare diseases like CLAs.[16,17] LLMs are sophisticated AI algorithms trained on vast amounts of web-sourced data that can understand and generate human-like text.[18] These LLM-based chatbots have the potential to expand access to trustworthy health information and support patient-engaged decision-making.[19] However, LLMs may also produce inaccurate information, potentially leading to confusion, frustration, or inappropriate medical decisions.[16,20] Accordingly, there is increasing emphasis on developing robust approaches for evaluating the quality of LLM-generated outputs.[19,21]

LLM-based chatbots usually generate responses in free-text form.[18] Unlike structured or exam-style outputs constrained by predefined formats (e.g. multiple-choice or checkbox options) that can be assessed against fixed gold-standard answers, free-text responses provide nuanced, contextualized, and personalized information, and thus require more sophisticated and flexible metrics to evaluate their quality. Although human evaluation remains the gold-standard for assessing answer quality and alignment with clinical expertise, it is time-consuming, resource-intensive, and difficult to scale consistently.[22,23] Given LLM-based chatbots’ capacity to rapidly generate large volumes of text, scalable and effective methods to evaluate these responses are urgently needed. Therefore, investigating the efficacy of automated methods for evaluating the quality of LLM-generated free-text responses is critical.

Most efforts to automatically evaluate free-text answers have relied on traditional natural language processing(NLP) metrics.[22] These include n-gram-based measures such as ROUGE, BLEU, and METEOR, as well as embedding-based semantic similarity metrics like BERTScore.[24–27] Originally developed for tasks such as text summarization and machine translation, these metrics assess the degree of lexical or semantic similarity between a generated response and a reference answer.[24–26] However, their effectiveness in evaluating nuanced answers to questions from rare disease patients remains uncertain.

An emerging alternative is LLM-based evaluation, also referred to as ***LLM-as-a-judge,*** which uses LLMs to assess the quality of another model’s output.[28,29] This approach offers the potential for consistent and scalable evaluation and has shown promising results in evaluating general NLP tasks, such as long-form summarization and open-domain QA.[21,30–34] However, their effectiveness for evaluating free-text responses to medical questions about rare diseases has not yet been investigated. To address this gap, we systematically assessed both traditional NLP metrics and LLM-based methods for evaluating LLM responses to patient questions about CLAs.

## MATERIALS AND METHODS

### CLA-QA dataset

We compiled a dataset consisting of 175 free-text responses generated by seven LLMs in response to 25 questions related to CLAs.

Question selection: A physician with expertise in CLAs (BAS) curated 25 questions about CLAs based on prior qualitative studies with patients and caregivers in this population, as well as “frequently asked questions” listed on the Lymphangiomatosis & Gorham’s Disease website, an advocacy group for patients with CLAs.[6,7,15] The question set included four subtypes of CLAs: Gorham-Stout disease, Generalized Lymphatic Anomaly, Kaposiform Lymphangiomatosis, Central Conducting Lymphatic Anomaly, and covered topics on disease definition, diagnosis, treatment, cause, family support, and mental health (see Table S1).

Answer-generation models: To generate responses for evaluation, we queried a leading proprietary model and a diverse set of open-source LLMs. The proprietary model, GPT-4, has demonstrated strong medical reasoning capabilities and high accuracy in addressing patient-facing rare disease questions.[35–37] The open-source models included LLaMA3.2-3.2B,[38] DeepSeek-R1-8B,[39] Phi-4-14B,[40] Mistral-7B,[41] Gemma 2-7B,[42] and Qwen 2-7B,[43] representing a range of model architectures and capabilities. Evaluating both proprietary and open-source models allowed for a broader assessment of model capabilities, including the potential of locally deployable LLMs for use in settings with cost, privacy, or security constraints.

Human annotation: Three domain-expert physicians specializing in CLAs (BAS, SCC, and KH) independently evaluated the accuracy of all 175 responses generated by the seven models. Annotators were blinded to the models that generated the answers. **Accuracy** was chosen as the primary evaluation criterion for evaluating answer quality, given its critical importance in medical chatbots. Accuracy was defined as the factual correctness of a response based on clinical expertise.[44] Ratings were assigned using a 5-point Likert scale, where 1 indicated a completely inaccurate answer and 5 indicated a completely accurate answer.

Reference answer selection: Many automatic evaluation methods, including traditional NLP sentence similarity metrics and LLM-based reference-guided scoring, require high-quality reference answers to enable consistent and scalable scoring of model outputs (described in detail later in MATERIALS AND METHODS-Automatic evaluation methods). Because expert-generated ideal answers are time-consuming to produce at scale, we instead identified the best-performing answer-generation model through expert evaluation and used its responses as reference answers for the reference-guided evaluation approaches. After selecting this model’s responses as the reference, we excluded them from subsequent automated evaluations.

### Automatic evaluation methods

#### NLP sentence similarity metrics

We used traditional NLP metrics to measure the similarity between generated responses and corresponding reference answers. Specifically, we evaluated responses using four commonly used metrics: ROUGE-L, BLEU, METEOR, and BERTScore.[24–27] Detailed descriptions of theses metrics are provided in Supplementary Section S1.These metrics served as proxies for answer quality by quantifying lexical or semantic similarity. Scores range from 0 and 1, with higher values reflecting greater similarity between the generated and reference answers.

#### LLM-based evaluation

We implemented two LLM-based evaluation methods to assess the quality of answers generated by different language models: reference-guided and reference-free scoring.

##### Reference-guided scoring

[45,46] The LLM evaluator was provided with a reference answer, along with the corresponding question and a model-generated response. The evaluator was then prompted to assign an accuracy score using a 5-point Likert scale, where higher scores indicate greater factual correctness. We prompted the LLMs to assess accuracy that aligned with the criterion used by physician annotators during human evaluation.

##### Reference-free scoring

[47] The LLM evaluator was only provided with the question and a model-generated response, without a reference answer. The evaluator was then asked to assign an accuracy score using the same 5-point Likert scale. To ensure consistency across evaluation conditions, we applied the same accuracy scoring criterion used in the human annotation and reference-guided evaluation.

We evaluated six LLM evaluators: GPT-4o,[48] GPT-4,[36] Gemma 3-27B,[49] DeepSeek-R1-14B,[39] Qwen 3-32B[50] and LLaMA3.3-70B.[38] Figure 1 provides an overview of the workflow for automating this evaluation of LLM-generated free-text responses. Open-source LLMs were deployed locally using Ollama on Washington University’s Research Infrastructure Services computing platform. Ollama enables efficient local execution of open-weight LLMs.[51]

**Figure 1.**
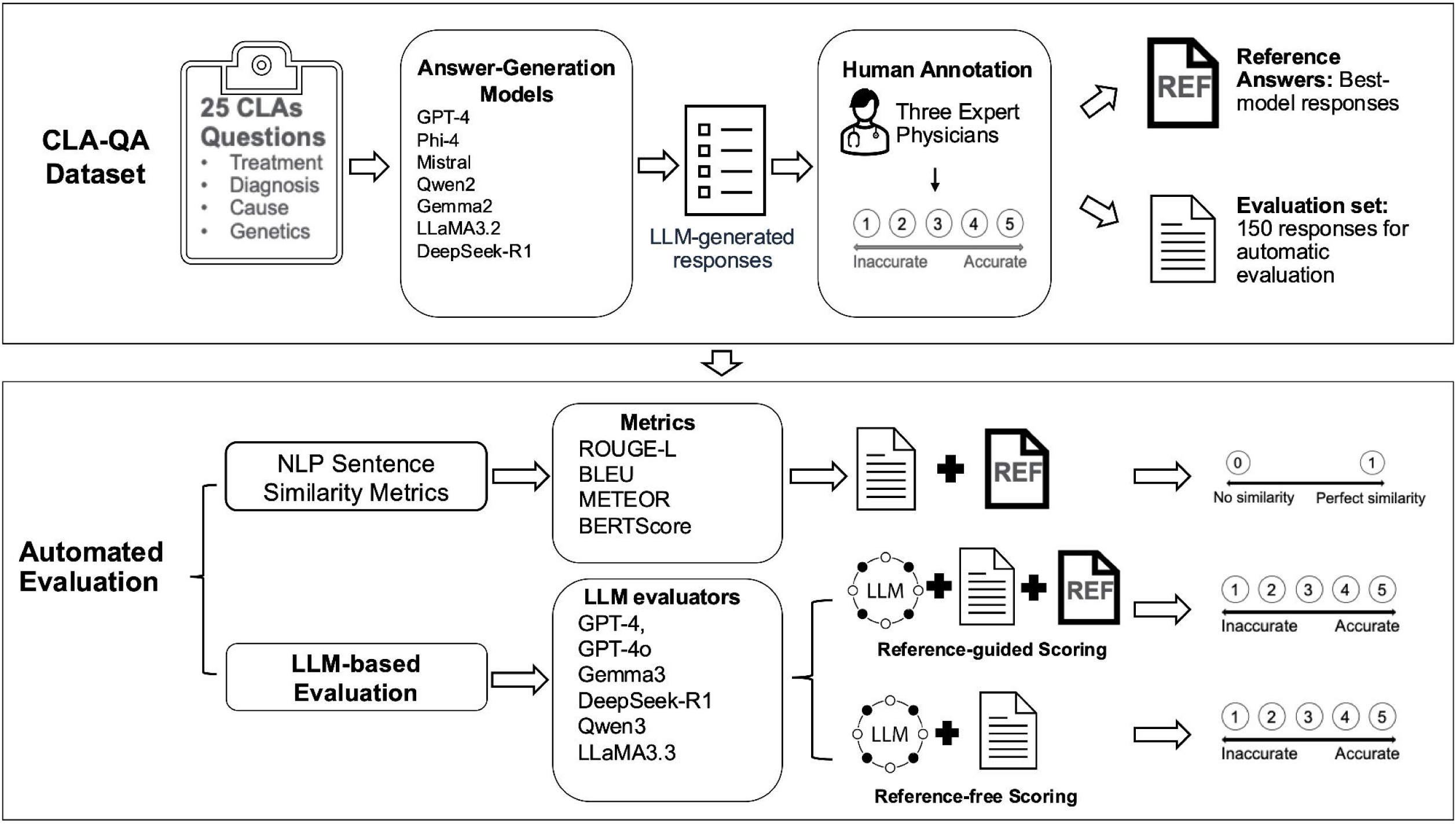
Overview of the workflow for automating evaluation of LLM-generated free-text responses.

These models were run with 32 GB RAM and a single NVIDIA A100 GPU. The GPT application programming interfaces (APIs) were hosted by Microsoft Azure’s OpenAI Service via a HIPAA-compliant subscription within WashU’s Azure tenant. The same prompts were used for every LLM evaluator. LLM evaluators’ temperatures were set to 0.

To assess the stability of LLM-based evaluation, we conducted 50 iterations of reference-guided scoring using LLM evaluators. For every response, scores were generated 50 times per evaluator. The variance across iterations was calculated for each response, and then the mean variance and standard deviation were computed for each LLM evaluator. To examine the impact of prompt design, we tested multiple prompt variants (see detailed prompts in Figure 2). Prompt A (Simple Instruction) directed the LLM to assign an accuracy score from 1 to 5, with no additional context or definitions. Prompt B (Basic Role + Task) framed the LLM evaluator as a CLA physician) and asked for an accuracy score. Prompt C (Definition Prompt) provided a definition of accuracy and evaluation criteria; we used this prompt as the ***default*** prompt in all subsequent analyses except for the prompt comparison. Prompt D (Definition + Rubric) included both a definition of accuracy and a detailed 1–5 scoring rubric for more consistent interpretation. We compared their performance by calculating the correlation between their assigned accuracy scores and physician-assigned scores. This analysis provided insight into the sensitivity of LLM evaluators to prompt structure and complexity.

**Figure 2.**
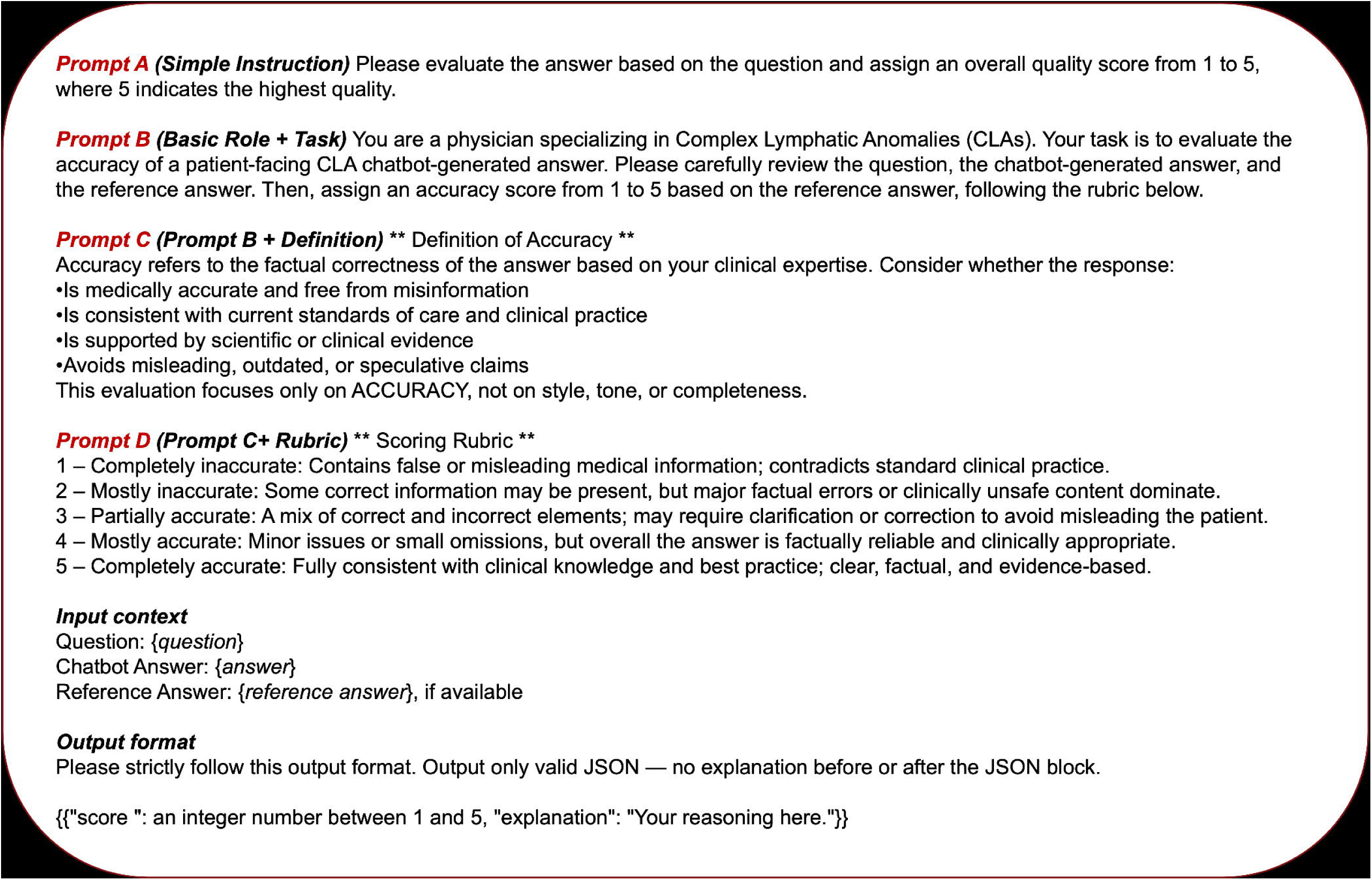
LLM-based Evaluation Prompt Design.

We analyzed two common sources of bias in LLM-based evaluation: verbosity bias and self-enhancement bias. Verbosity bias, also known as length bias, refers to the tendency of LLM evaluators to favor responses of a particular length, often showing a preference for more verbose outputs.[52] We tested this by computing the Spearman correlation between answer length and accuracy scores assigned by physicians and LLM evaluators under the reference-guided setting. Self-enhancement bias refers to the tendency of LLM evaluators to favor responses generated by themselves or by models similar to themselves.[21] We assessed self-enhancement bias by comparing how each LLM evaluator scored responses generated by its corresponding model family.

### Statistical analysis

We assessed inter-rater reliability among the three expert physician reviewers using the intraclass correlation coefficient (ICC).[53,54] To evaluate response-level associations, we used Spearman’s rank correlation coefficient (ρ) to assess the relationship between physician-assigned scores, traditional NLP metric scores, and LLM-assigned scores.[55] We interpreted correlation strength using the following thresholds: 0.00–0.10 negligible, 0.10–0.39 weak, 0.40–0.69 moderate, 0.70–0.89 strong, and 0.90–1.00 very strong correlations.[56] At the answer-generation model level, we calculated Kendall’s τ to assess correlations between the average physician-assigned scores and LLM-assigned accuracy scores.[57] We used descriptive statistics to summarize average NLP metric values, LLM-assigned accuracy scores, and inference time for each evaluator. To test differences in model scores across three or more related groups, we applied the Friedman test, using a significance threshold of p < 0.05.[58]

To further evaluate the binary discrimination ability of LLM-based scoring methods, we recoded physician-assigned and LLM-assigned scores into binary labels, with scores of 1–3 categorized as low accuracy and 4–5 as high accuracy. We then calculated Phi correlation coefficients (φ) between the binary labels derived from physician-assigned scores and those derived from LLM-assigned scores, thereby assessing each LLM evaluator’s ability to distinguish between low- and high-quality responses.[59] All statistical analyses were conducted using Python 3.12.

## RESULTS

### Human annotation results

Inter-rater reliability among three expert physicians was high (ICC=0.852), indicating substantial agreement across raters. In all subsequent analyses, “**physician-assigned score**” refers to the mean accuracy score from three expert physicians per response.

To enable consistent and scalable evaluation of LLM-generated free-text responses, we first identified a high-quality set of reference answers. Expert physicians rated GPT-4’s responses highest in accuracy (mean score 4.77, SD=0.25); therefore, we selected these responses as the reference set for the reference-guided evaluation methods (see Figure 3 and Table S2). After designating GPT-4’s responses as the reference, we excluded them from subsequent evaluation of free-text responses using automated methods. Among the remaining answer-generation models evaluated, Phi-4 achieved the highest average physician-assigned score of 3.53 (SD=1.25), while DeepSeek-R1-8B received the lowest average score (2.64, SD=1.33). The Friedman test showed a significant difference in physician-assigned scores across the six evaluated models (p=0.005).

**Figure 3.**
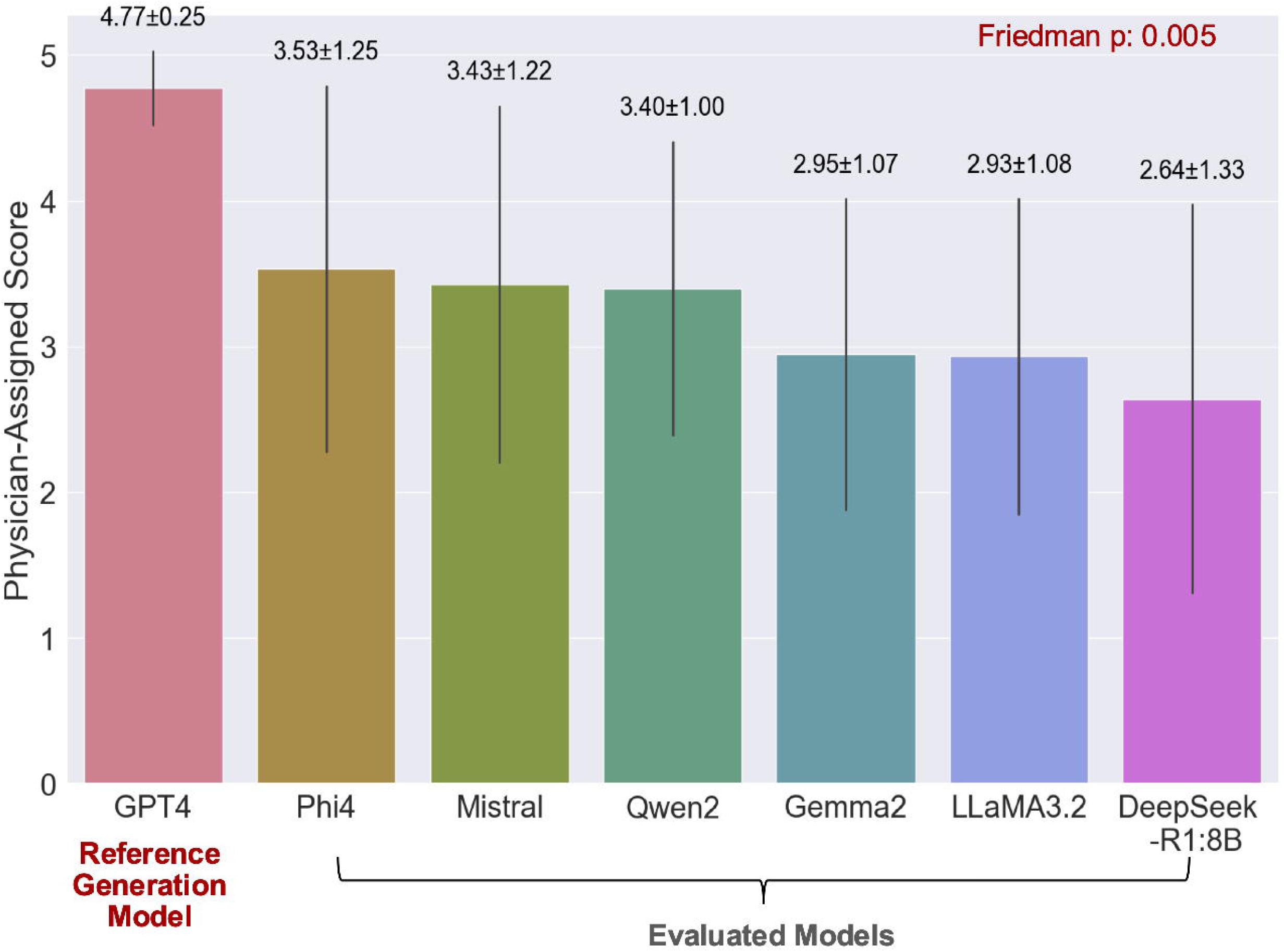
Comparison of LLMs’ accuracy on CLA-QA based on physician-assigned score. Bars show the mean physician-assigned scores over 25 answers for each answer-generation model. Error bars represent standard deviations, and the mean ± SD values are displayed above each bar for reference.

### Effectiveness of automated evaluation methods

#### Reference-based evaluation of NLP metrics and LLM evaluators

We computed NLP sentence similarity metrics and conducted reference-guided scoring with LLM evaluators using reference answers. To assess their alignment with expert judgment, we calculated the Spearman correlation between the resulting automatic evaluation scores and physician-assigned scores.

Among four NLP sentence similarity metrics assessed, METEOR had the lowest correlation with physician-assigned scores (Spearman ρ=0.240), while BERTScore had the strongest correlation (ρ=0.406). Among LLM evaluators, GPT-4 achieved the highest correlation with physician-assigned scores (ρ=0.758), followed closely by GPT-4o (ρ=0.727). All four open-source LLM evaluators demonstrated moderate correlations with physician-assigned scores, ranging from 0.506 (DeepSeek-R1-14B) to 0.608 (Qwen3) (Table 1).

**Table 1.** Correlation of NLP sentence similarity metrics and reference-guided LLM-based evaluation with physician assessment.

|  | Correlation with Physician-Assigned Score |  |
| --- | --- | --- |
| | Spearman $\rho$ | p value |
| *NLP Sentence Similarity Metrics* |  |  |
| **ROUGE-L** | 0.325 | $p < 0.001$ |
| **BLEU** | 0.361 | $p < 0.001$ |
| **METEOR** | 0.240 | 0.003 |
| **BERTScore** | 0.406 | $p < 0.001$ |
| *LLM evaluator using Reference-guided Scoring* |  |  |
| **GPT-4 Evaluator** | **0.758** | $p < 0.001$ |
| **GPT-4o Evaluator** | 0.727 | p < 0.001 |
| **Gemma3-27B Evaluator** | 0.552 | p < 0.001 |
| **DeepSeek-R1-14B Evaluator** | 0.506 | p < 0.001 |
| **Qwen3-32B Evaluator** | 0.608 | p < 0.001 |
| **LLaMA3.3-70B Evaluator** | 0.596 | p < 0.001 |

\* The highest score is shown in bold.

To further examine how these evaluation metrics differentiate free-text answer quality across models, we compared the mean accuracy scores of responses generated by different models using both classic NLP sentence similarity metrics with LLM-based reference-guided evaluators (Table 2). Across the six answer-generation models, NLP sentence similarity metrics exhibited relatively narrow score ranges across models. All NLP metrics captured some performance differences, they demonstrated statistically significant variation in model scores based on the Friedman test (p<0.001), indicating their ability to differentiate among the evaluated models, with at least one model performing significantly better than the others. However, the resulting performance rankings were not aligned with physician-assigned scores. For instance, the DeepSeek-R1-8B model received the lowest average score from expert reviewers yet was rated relatively highly by several NLP metrics.

**Table 2.**
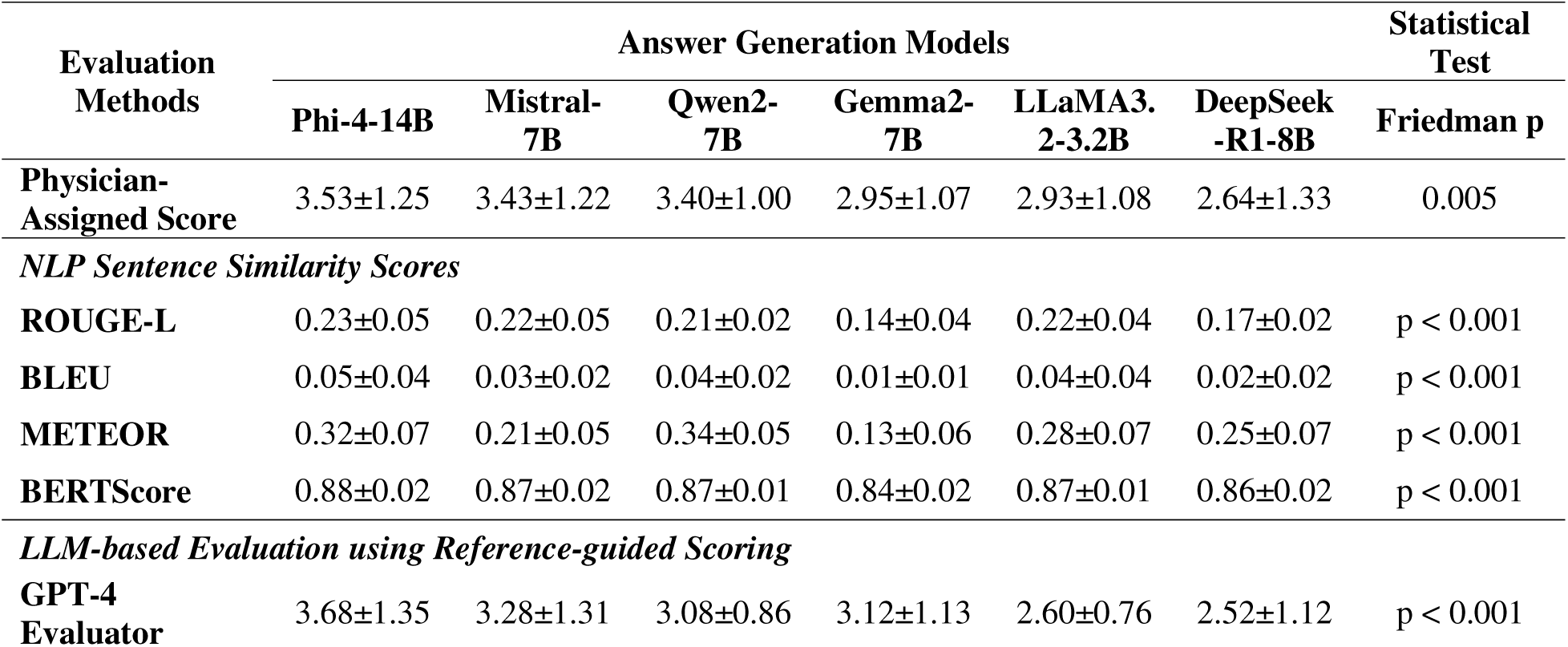
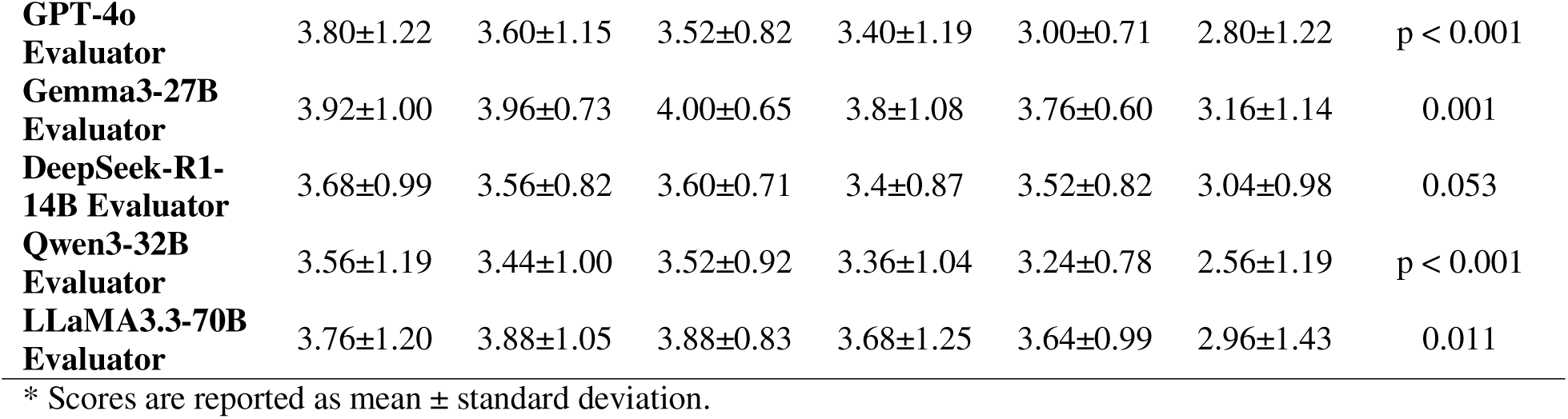
Performance of answer-generation models evaluated by NLP metrics and LLM-based reference-guided scoring.

| Evaluation Methods | Answer Generation Models |  |  |  |  |  | Statistical Test |
| --- | --- | --- | --- | --- | --- | --- | --- |
|  | Phi-4-14B | Mistral-7B | Qwen2-7B | Gemma2-7B | LLaMA3.2-3.2B | DeepSeek-R1-8B | Friedman p |
| **Physician-Assigned Score** | 3.53±1.25 | 3.43±1.22 | 3.40±1.00 | 2.95±1.07 | 2.93±1.08 | 2.64±1.33 | 0.005 |
| *NLP Sentence Similarity Scores* |  |  |  |  |  |  |  |
| **ROUGE-L** | 0.23±0.05 | 0.22±0.05 | 0.21±0.02 | 0.14±0.04 | 0.22±0.04 | 0.17±0.02 | p < 0.001 |
| **BLEU** | 0.05±0.04 | 0.03±0.02 | 0.04±0.02 | 0.01±0.01 | 0.04±0.04 | 0.02±0.02 | p < 0.001 |
| **METEOR** | 0.32±0.07 | 0.21±0.05 | 0.34±0.05 | 0.13±0.06 | 0.28±0.07 | 0.25±0.07 | p < 0.001 |
| **BERTScore** | 0.88±0.02 | 0.87±0.02 | 0.87±0.01 | 0.84±0.02 | 0.87±0.01 | 0.86±0.02 | p < 0.001 |
| *LLM-based Evaluation using Reference-guided Scoring* |  |  |  |  |  |  |  |
| **GPT-4 Evaluator** | 3.68±1.35 | 3.28±1.31 | 3.08±0.86 | 3.12±1.13 | 2.60±0.76 | 2.52±1.12 | p < 0.001 |
| **GPT-4o Evaluator** | 3.80±1.22 | 3.60±1.15 | 3.52±0.82 | 3.40±1.19 | 3.00±0.71 | 2.80±1.22 | p < 0.001 |
| **Gemma3-27B Evaluator** | 3.92±1.00 | 3.96±0.73 | 4.00±0.65 | 3.8±1.08 | 3.76±0.60 | 3.16±1.14 | 0.001 |
| **DeepSeek-R1-14B Evaluator** | 3.68±0.99 | 3.56±0.82 | 3.60±0.71 | 3.4±0.87 | 3.52±0.82 | 3.04±0.98 | 0.053 |
| **Qwen3-32B Evaluator** | 3.56±1.19 | 3.44±1.00 | 3.52±0.92 | 3.36±1.04 | 3.24±0.78 | 2.56±1.19 | p < 0.001 |
| **LLaMA3.3-70B Evaluator** | 3.76±1.20 | 3.88±1.05 | 3.88±0.83 | 3.68±1.25 | 3.64±0.99 | 2.96±1.43 | 0.011 |

\* Scores are reported as mean ± standard deviation.

In comparison, all LLM evaluators exhibited variation in scoring answer-generation models. The GPT-4 evaluator assigned the highest average score of 3.68 (SD=1.35) to Phi-4, which was aligned closely with its highest physician-assigned average score of 3.53 (SD=1.25). Similarly, the GPT-4o evaluator assigned the highest score for Phi-4 (3.80, SD=1.22), followed by 3.60 (SD=1.15) for Mistral. Overall, Figure 4 illustrates that GPT evaluators closely align with physician-assigned scores, whereas NLP sentence similarity metrics diverge substantially, in terms of relative model ranking. Except for the DeepSeek-R1-14B evaluator (Friedman test, p=0.053), all other LLM evaluators showed statistically significant differences in scores across answer-generation models (p<0.05), indicating their ability to distinguish model performance.

**Figure 4.**
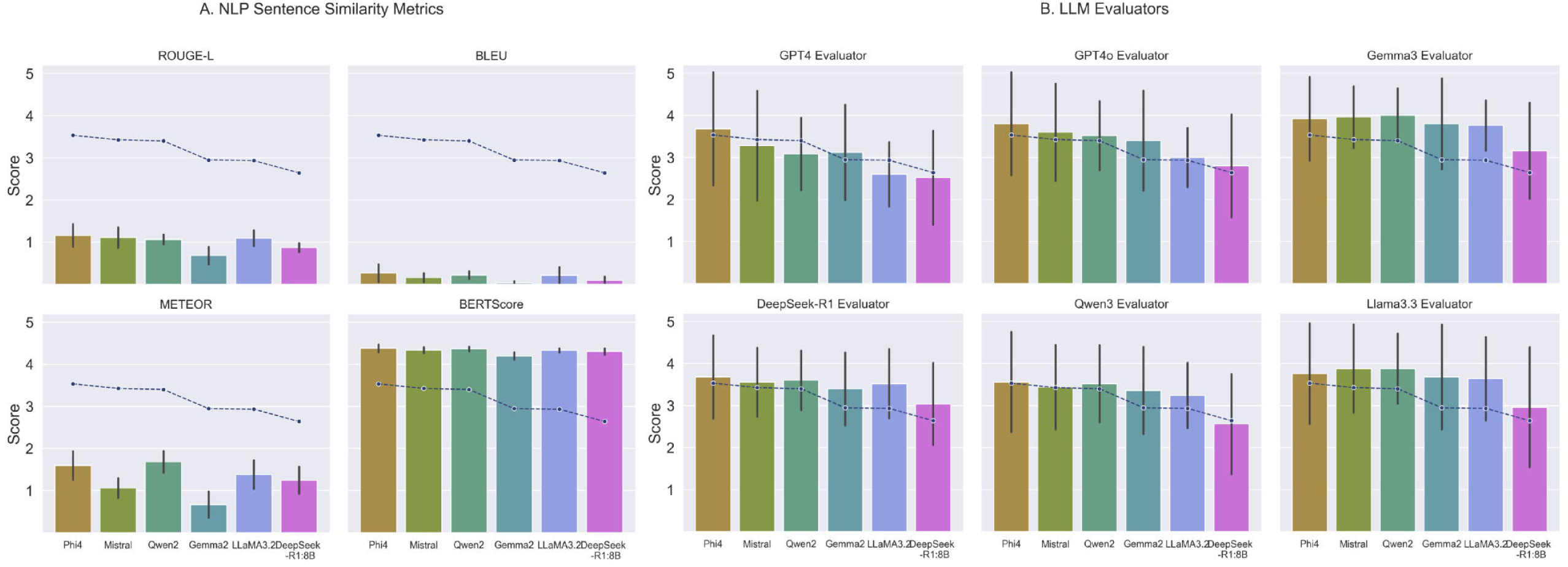
Comparison of answer-generation models performance using physician-assigned scores, NLP sentence similarity metrics and reference-guided LLM-based evaluations. The dashed line represents the mean physician-assigned scores, indicating the relative performance ranking of the models. Bar plots show the mean accuracy scores for each model across different automated evaluation methods, with error bars representing standard deviations. The NLP scores were rescaled from [0–1] to [0–5] to align with the 5-point accuracy scale in the visualization.

#### Comparison across LLM-based evaluation methods

##### Reference-guided versus reference-free scoring

We next further characterized agreement between automated LLM evaluators and physician-assigned scores, both for reference-free scoring and reference-guided scoring.

When evaluating each response produced by the answer-generation models, GPT4 and GPT4o-assigned scores correlated more strongly with physician-assigned scores under reference-guided methods (ρ=0.758 and 0.727, respectively) than under reference-free methods (ρ=0.661 and 0.662, respectively). All other models also showed weaker correlations under reference-free compared to reference-guided methods, except for Qwen3, which demonstrated a slight increase in correlation with physician-assigned scores, from ρ=0.608 to ρ=0.614 (Table 3).

**Table 3.** Correlation of LLM-based evaluation methods with physician-assigned scores.

| ***Response Level (150 responses)*** |  |  |  |  |
| --- | --- | --- | --- | --- |
| Evaluation Method | Reference-guided Scoring |  | Reference-free Scoring |  |
| *LLM evaluators* | Spearman $\rho^1$ | Binary Correlation<sup>2</sup><br>$\phi$ | Spearman $\rho$ | Binary Correlation<br>$\phi$ |
| GPT-4 Evaluator | 0.758 | 0.508 | 0.690 | 0.501 |
| GPT-4o Evaluator | 0.727 | 0.500 | 0.700 | 0.415 |
| Gemma3-27B Evaluator | 0.552 | 0.314 | 0.388 | 0.261 |
| DeepSeek-R1-14B Evaluator | 0.506 | 0.304 | 0.183 | 0.131 |
| Qwen3-32B Evaluator | 0.608 | 0.365 | 0.614 | 0.419 |
| LLaMA3.3-70B Evaluator | 0.596 | 0.349 | 0.409 | 0.179 |
| ***Answer Generation Model Level (6 models)*** |  |  |  |  |
| Evaluation Method | Reference-guided Scoring |  | Reference-free Scoring |  |
| *LLM evaluators* | Kendall's $\tau^1$ | p value | Kendall's $\tau$ | p value |
| GPT-4 Evaluator | 0.867 | 0.02 | 0.867 | 0.02 |
| GPT-4o Evaluator | 1.000 | 0.003 | 0.867 | 0.017 |
| Gemma3-27B Evaluator | 0.600 | 0.136 | 0.690 | 0.136 |
| DeepSeek-R1-14B Evaluator | 0.733 | 0.056 | 0.690 | 0.136 |
| Qwen3-32B Evaluator | 0.867 | 0.017 | 0.966 | 0.017 |
| LLaMA3.3-70B Evaluator | 0.690 | 0.056 | 0.552 | 0.056 |

After recoding the 5-point scores into binary categories — low accuracy (1-3) and high accuracy (4-5) — both LLM-based reference-guided and reference-free methods showed reduced alignment with physician-assigned scores as measured by the Phi coefficient. Among the LLM evaluators, GPT-4o-assigned scores exhibited the strongest alignment with physician-assigned scores (φ=0.508, moderate) under the reference-guided approach, whereas the DeepSeek-R1-14B evaluator showed the weakest correlation, indicating limited ability to distinguish between high- and low-quality responses in binary categorization.

1. Binary Phi correlation was calculated by recoding the 5-point accuracy scale into two categories: low accuracy (scores 1-3) and high accuracy (scores 4-5).
2. All response-level correlation results were statistically significant (p < 0.05).

To assess the extent to which evaluators ranked each model’s overall performance similarly to physicians, we computed Kendall’s τ between the average physician-assigned scores and LLM-assigned scores at the model level. GPT-4 and GPT-4o evaluators demonstrated strong rank correlation with physicians’ judgments, with Kendall’s τ ranging from 0.867 to 1.0. Among open-source LLM evaluators, Qwen3 evaluator exhibited the strongest alignment with physician, achieving τ = 0.867 under reference-guided scoring and τ = 0.966 under reference-free scoring. Figure 5 illustrates the comparative results of both LLM-based evaluation methods across all LLM evaluators. Under the reference-free setting, LLM evaluators’ scores showed greater deviation from physician-assigned scores than under reference-guided scoring. Supplementary Table S3 reports the mean scores for each answer-generation model under the reference-free scoring setting.

**Figure 5.**
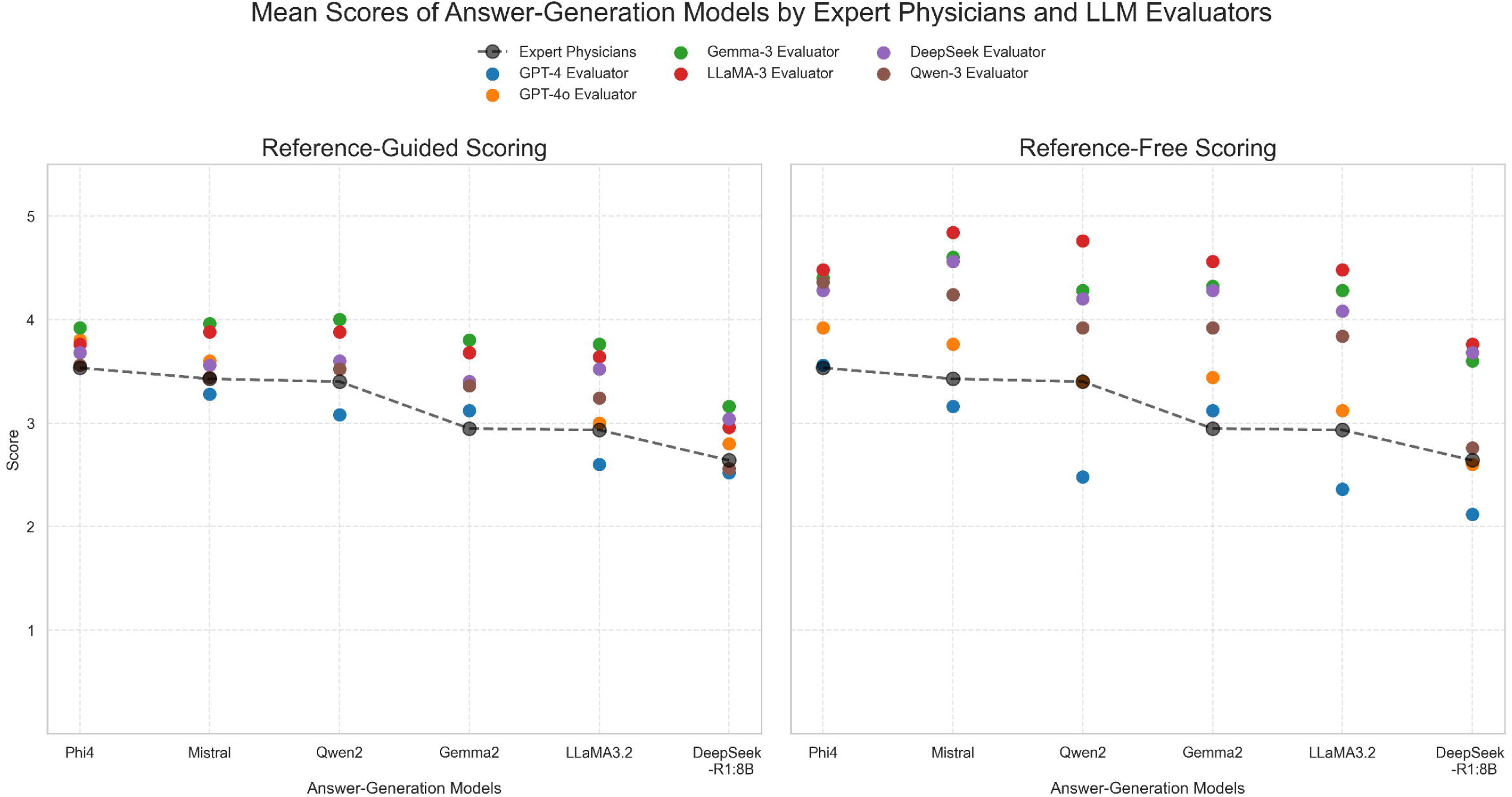
Comparison of reference-guided versus reference-free scores assigned by LLM evaluators across answer-generation models. Mean accuracy scores assigned to each answer-generation model by expert physicians and LLM evaluators using reference-guided (left) and reference-free (right) scoring methods. Scores are reported on a 5-point Likert scale. Each point represents the mean score assigned by either expert physicians, or an LLM evaluator for a given answer generation model, using the reference-guided scoring method. Black dashed lines represent physician-assigned scores; colored markers denote different LLM evaluators.

### Practical considerations in LLM-based evaluation

#### Stability of LLM-based evaluations

To assess the stability of LLM evaluators, we conducted 50 replicates of reference-guided scoring. The mean variance across all responses was 0.048 (SD=0.26) for the GPT-4o evaluator and 0.009 (SD=0.037) for the GPT-4 evaluator, indicating high stability. A notable exception was observed for a GPT-4-generated response regarding Kaposiform Lymphangiomatosis (KLA)’s hereditary risk that received a physician-assigned score of 5. For this response, the GPT-4o evaluator’s scores varied considerably across runs: 52% were rated as 5, 14% as 3, and 34% as 1. This highlighted a potential limitation in scoring consistency when evaluating clinically complex answers. Meanwhile, four open-source LLM evaluators demonstrated perfect scoring consistency (variance=0) across 50 runs, due to deterministic outputs under a fixed temperature of zero.

#### Impact of prompt design

To examine the sensitivity of LLM evaluators to prompt structure and complexity, we tested four prompt variants and assessed their impact on LLM-based evaluation performance using Spearman correlation coefficients (Figure 6). For GPT-4 and GPT-4o evaluators, prompt design had minimal influence on performance: correlations with physician-assigned scores remained consistently high across prompts (ρ=0.699–0.761) under the reference-guided setting and moderately high under the reference-free setting (ρ=0.667–0.731). By comparison, open-source LLM evaluators exhibited greater variability in correlation with physician-assigned scores depending on prompt design, particularly in the reference-free setting. Despite some improvement with more structured prompts, their correlations remained in the moderate or weak range.

**Figure 6.**
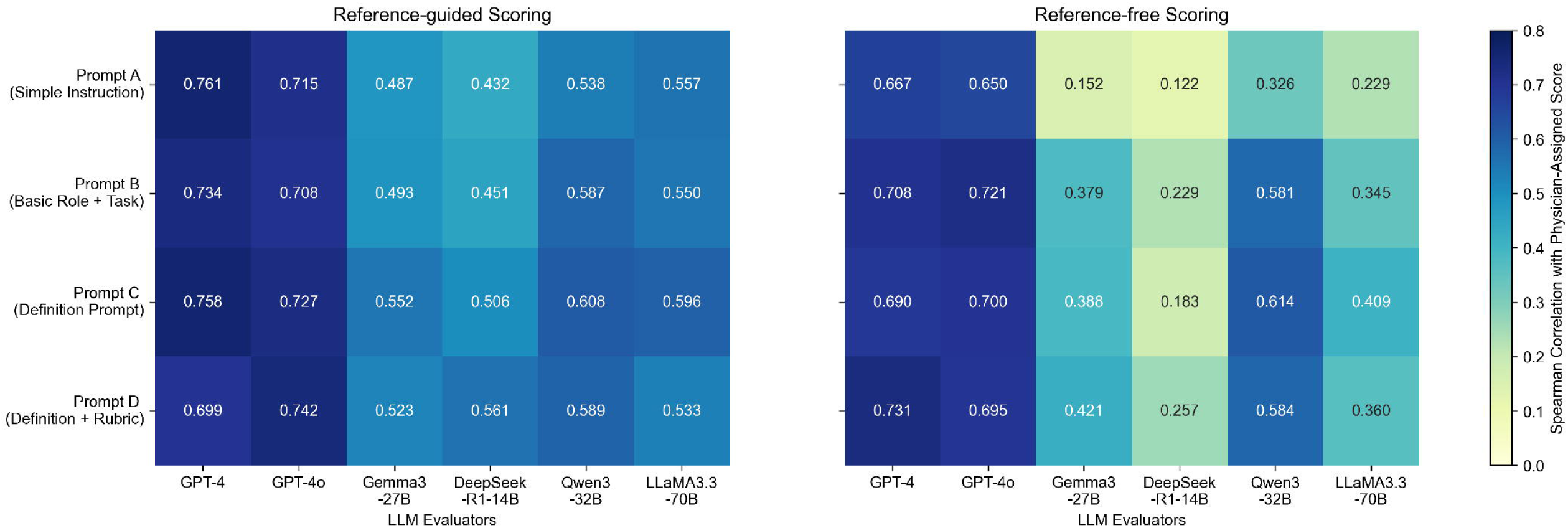
Effect of prompt variants on LLM evaluator performance measured by Spearman correlation with Physician-assigned scores. Heatmaps showing that GPT-4 and GPT-4o evaluators maintain consistently high correlations with physician-assigned scores across prompts, while open-source LLM evaluators vary more widely and show weaker correlations, especially in the reference-free setting.

#### Inference time and computation cost

To evaluate the feasibility of LLM-based evaluation, we recorded inference time and computation cost. GPT-4 and GPT-4o evaluators were the most time-efficient, averaging 1.68 (SD=0.66) and 1.51 (SD=0.57) seconds per responses, respectively. Open-source LLM evaluators such as Qwen3 and LLaMA3.3 required over 30 and 200 seconds per response (Table S4). Regarding computational cost, GPT-4 and GPT-4o evaluators incurred estimated per-response costs of (8.3±1.4)×10 ³ USD and (2.30±0.37)×10 ³ USD, respectively. Open-source LLM evaluators were run locally using Ollama and did not incur additional per-inference costs.

#### Bias in LLM-based evaluation

##### Verbosity Bias

To investigate whether answer length influenced scores, we calculated the Spearman correlation between LLM-assigned scores and answer length. Correlations ranged from 0.025 (GPT-4) to 0.162 (DeepSeek-R1-14B), suggesting weak verbosity effects (Table S5). The correlation between physician-assigned scores and answer length was 0.167, indicating no significant association.

##### Self-enhanced bias

We examined whether LLM evaluators favored their own model family under reference-guided scoring (Table 2). The Gemma3 evaluator assigned higher scores to Gemma2 responses (3.80, SD=1.08), which exceeded the physician-assigned rating 2.95 (SD=1.07). DeepSeek-R1-14B rated its own model’s responses at 3.04(SD=0.98), compared to a lower physician-assigned score of 2.64 (SD=1.33). Similarly, the LLaMA3.3 evaluator gave a score of 3.64 (SD=0.87) to LLaMA3.2 responses, also higher than the physician score of 2.93(SD=1.08). Qwen3 evaluator did assigned a score of 3.52 to Qwen2 responses, close to the physician-assigned score of 3.40.

However, while these LLM evaluators assigned higher scores to their own models compared to physician ratings, they also consistently gave elevated scores across all models, indicating general score inflation rather than targeted self-favoritism.

## DISCUSSION

With the growing need to develop LLM-based chatbots for rare disease patients and families, we must address the challenge of assessing the quality of large volumes of free-text responses generated by these systems. Because human evaluation is not feasible at scale, automation becomes essential. Automating the evaluation of LLM-generated responses is a critical yet challenging task, given the clinical importance of ensuring nuanced and accurate information to meet the needs of rare disease patients and families. In this study, we systematically compared traditional NLP sentence similarity metrics with LLM-based evaluation methods to assess their effectiveness in approximating expert physicians’ judgment of the accuracy of LLM output. Our findings provide valuable insights into the performance, reliability, and limitations of automated evaluation methods.

Our results showed that LLM-based evaluation outperformed traditional NLP metrics, with LLM-assigned scores achieving higher correlations with physician-assigned scores. Compared with LLM-based evaluation results, ROUGE-L, BLEU, METEOR and BERTScore exhibited lower correlations at response-level with physician-assigned scores, highlighting their limitations in evaluating LLM-generated free-text responses to rare disease questions. While they were able to detect some performance differences across answer-generation models, as indicated by the Friedman test (p < 0.001), their resulting rankings did not consistently align with expert physician evaluations. This may be attributed to their underlying rationale: n-gram-based metrics mainly assess lexical similarity and phrase overlap, while semantic similarity metrics like BERTScore rely on contextual embeddings. The quality and domain relevance of embedding models significantly affect their performance; however, embedding models are typically pre-trained and fixed, limiting their flexibility and adaptability to specialized medical domains.[60,61]

Another limitation of traditional NLP metrics is their scalability, as they require a reference answer to generate scores, making their effectiveness highly dependent on the availability of large numbers of high-quality reference answers, which constrains scalability. In contrast, LLM-based evaluation methods, particularly structured scoring methods, can be applied in both reference-guided and reference-free settings, offering greater flexibility and demonstrating better alignment with expert physicians’ judgment. GPT-4 and GPT-4o evaluators consistently demonstrated the strongest response-level correlation with physician-assigned scores across both reference-guided and reference-free scoring strategies. Their rankings closely mirrored expert physicians’ rankings of model performance, supporting their reliability for model-level assessment. Among open-source models, Qwen3 demonstrated relatively stronger correlations with physician-assigned scores compared to Gemma3, DeepSeek-R1-14B, and LLaMA3.3.

Compared to reference-guided scoring, reference-free evaluation offered greater flexibility by not requiring reference answers but showed lower correlations with physician-assigned scores using LLM evaluators. Additionally, reference-free scoring exhibited wider scoring ranges and greater variability, particularly with open-source LLMs. Both scoring methods demonstrated reduced alignment with physician annotations in binary classification, underscoring the greater reliability of 5-point scoring rubrics.

Stability analyses showed low or no variance in repeated LLM-based evaluation under a fixed temperature setting of zero, supporting their reproducibility. While GPT-4 and GPT-4o evaluators were relatively robust to prompt variation, open-source models showed greater sensitivity to prompt design, particularly in reference-free settings. These findings underscored the need for further research to identify optimal prompt designs for evaluation tasks. Our bias analysis indicated minimal verbosity bias, with weak correlations between response length and evaluation scores across all models. Gemma3, DeepSeek-R1-14B, LLaMA3.3 and Qwen3 evaluators tended to assign higher scores to responses generated by their own model families, though this pattern appeared to reflect general score inflation rather than targeted self-enhancement. Further research is needed to investigate these biases in more depth.

These findings have several novel insights on the strengths and limitations of automated evaluation methods for assessing free-text responses. The high correlation with physician-assigned scores and short inference time of LLM-based evaluation, especially proprietary models like the GPT series, highlighted their potential for enabling large-scale, automated evaluation systems that approximate physicians’ judgment. Among the LLM-based evaluation methods assessed, those incorporating reference answers showed stronger alignment with physician-assigned scores, indicating that reference-guided scoring was the most effective approach for approximating physicians’ judgment. These automated evaluation methods could substantially reduce the resource burden of human review for LLM-based rare disease chatbots outputs.

Despite these promising results for LLM-based evaluation, several limitations must be acknowledged. First, the focus on CLAs may limit generalizability to other rare diseases or broader contexts. Second, the physician annotations from three board-certified clinical experts, while robust, inherently involved subjective judgments, and thus the use of aggregated expert evaluations may not eliminate bias or variability. Third, we assessed only accuracy, without considering other attributes such as clarity or lay-friendliness, which are essential for patient-facing chatbots. Finally, we used GPT-4-generated answers as reference answers for reference-guided evaluations. Although GPT-4 outputs are rated highly by physicians, they may introduce subtle biases or reinforce model-specific phrasing, potentially favoring similar models. Future research should test robustness across diverse prompts, question types, and diseases, and explore methods to enhance open-source models to achieve near expert-level evaluations in rare disease contexts through domain adaptation and retrieval-augmented techniques. Additionally, future work could examine the effectiveness of combining multiple automated methods, such as integrating sentence similarity metrics with LLM-based evaluation, to improve overall evaluation accuracy and reliability.

## CONCLUSION

To the best of our knowledge, this study is the first to demonstrate the application of automated LLM-based evaluation methods for free-text responses in the rare disease domain. Our results highlighted the potential of LLM-based evaluation, particularly GPT series models, as scalable, automated alternatives to human review in evaluating LLM-generated responses to patient questions about rare diseases. While LLM-based evaluation approaches show promising results, they must be carefully designed, validated, and continuously monitored to ensure their outputs remain clinically relevant and aligned with human judgment. Future work should further explore LLM-based evaluation robustness and generalizability across diverse rare disease domains.

## Supporting information

Supplementary Materials, Table S1, Section S1, Table S2, Table S3, Table S4, Table S5

## ACKNOWLEDGMENTS

This work was supported in part by a pilot grant from the Orphan Disease Center at the University of Pennsylvania through the Million Dollar Bike Ride program, and a pilot grant from Pedal the Cause at Siteman Cancer Center and Siteman Kids at St. Louis Children’s Hospital.

## DATA AVAILABILITY

The CLA-QA dataset, consisting of 25 common CLA patient questions, 175 responses generated by seven LLMs, and accuracy annotations from three expert physicians, is available in the Washington University institutional repository at https://data.library.wustl.edu/record/108301?v=tab.

## AUTHOR CONTRIBUTIONS

Min Zhao (Conceptualization, Data Curation, Formal analysis, Methodology, Investigation, Writing—original draft, Writing—review & editing) Inez Oh (Conceptualization, Methodology, Investigation, Writing – review & editing), Aditi Gupta (Conceptualization, Methodology, Investigation, Writing – review & editing), Sally Cohen-Cutler (Data Curation, Funding acquisition, Writing – review & editing), Kathryn M Harmoney (Data Curation, Writing – review & editing), Albert M. Lai (Conceptualization, Methodology, Investigation, Supervision, Writing – review & editing), Bryan A. Sisk (Conceptualization, Methodology, Investigation, Supervision, Data Curation, Funding acquisition, Writing – review & editing)

