## Supplementary Materials, Table S1, Section S1, Table S2, Table S3, Table S4, Table S5 for "Automating Evaluation of LLM-generated Responses to Patient Questions about Rare Diseases"

### Table S1. CLA-QA dataset question list

**CLA Questions**

1. What is Gorham Stout Disease?

2. Is Gorham Stout Disease hereditary?

3. What can make Gorham Stout Disease become active? Trauma, infection, puberty, pregnancy,…

4. Why is having surgery or a biopsy risky for a Gorham-Stout Disease patient? Why can they not remove the lymphatic mass?

5. Does generalized lymphatic anomaly seem to stay localized to the parts of the body that were originally affected or does it spread?

6. What are the reported side effects of sirolimus for generalized lymphatic anomaly?

7. Is it dangerous to have children if I have kaposiform lymphangiomatosis?

8. Why do they have to use drug combinations for a kaposiform lymphangiomatosis patient?

9. Does Lymphatic Drainage Massage work for central conducting lymphatic anomaly?

10. Low fat diets - do they work for central conducting lymphatic anomaly?

11. What do lymphatics do in your body?

12. What type of imaging do I need for Gorham Stout Disease? How often do I need to get imaging?

13. Does Lymphatic Drainage Massage work for Gorham-Stout Disease?

14. How long can/should a Gorham-Stout Disease patient be on sirolimus? Can it be used for life? Should I/my child be monitored whilst on this drug? What does that look like?

15. How is generalized lymphatic anomaly diagnosed?

16. What genes cause generalized lymphatic anomaly? Does generalized lymphatic anomaly run in families?

17. What are the symptoms of generalized lymphatic anomaly?

18. What types of doctors take care of patients with Kaposiform lymphangiomatosis?

19. What type of imaging do I need for kaposiform lymphangiomatosis? How often do I need to get imaging?

20. What are the symptoms of kaposiform lymphangiomatosis?

21. Why do we get so much pain with kaposifor lymphangiomatosis?

22. Vaccine - Is it safe for me/my child to get the COVID vaccine given our diagnosis of kaposiform lymphangiomatosis? Should we receive the flu and pneumonia vaccine? Can we have live vaccines? Could a vaccine trigger the disease to become active?

23. What are the symptoms of central conducting lymphatic anomaly?

24. How long can/should a central conducting lymphatic anomaly patient be on sirolimus? Can it be used for life? Should I/my child be monitored whilst on this drug? What does that look like?

25. Should I be taking supplements if I have central conducting lymphatic anomaly? If so, what?

### Section S1. Description of NLP sentence similarity metrics

*ROUGE* ***(Recall-Oriented Understudy for Gisting Evaluation)***:[24] ROUGE measures the overlap of n-grams, word sequences, and word pairs between generated and reference answers. In this study, we used ROUGE-L, a variant that calculates similarity based on the longest common subsequence and accounts for sentence-level structural similarity.

*BLEU* ***(Bilingual Evaluation Understudy)***:**[25]** Originally developed for machine translation, BLEU calculates the precision of n-grams (typically up to 4-grams) in the generated answers that also appear in the reference answer. It incorporates a brevity penalty to discourage overly short outputs, thereby favoring concise and accurate responses.

*METEOR* ***(Metric for Evaluation of Translation with Explicit ORdering)***:**[26]** METEOR improves on BLEU by incorporating both precision and recall of unigrams and by aligning words using stemming, synonyms, and paraphrase matching.

*BERTScore*:[27] A semantic similarity metric that uses contextual embeddings from pretrained BERT models to compare token-level similarity between generated and reference responses.[62] Unlike surface-level overlap metrics, BERTScore can capture deeper semantic meaning and paraphrasing.

### Table S2. Average accuracy scores assigned by expert physician reviewers.

| **Answer Generation Model** | **Reviewer-BAS** | **Reviewer-KH** | **Reviewer-SCC** | **Physician-Assigned Score (Mean)** |
| --- | --- | --- | --- | --- |
| **GPT-4** | 4.64±0.57 | 4.72±0.54 | 4.96±0.2 | 4.77±0.25 |
| **Phi-4-14B** | 3.20±1.41 | 3.76±1.30 | 3.64±1.58 | 3.53±1.25 |
| **Mistral-7B** | 3.00±1.41 | 3.60±1.22 | 3.68±1.44 | 3.43±1.22 |
| **Qwen2-7B** | 2.92±1.32 | 3.68±0.99 | 3.60±1.44 | 3.40±1.00 |
| **Gemma2-7B** | 2.48±1.29 | 3.44±1.19 | 2.92±1.32 | 2.95±1.07 |
| **LLaMA3.2-3.2B** | 2.48±1.36 | 3.24±1.13 | 3.08±1.26 | 2.93±1.08 |
| **DeepSeek-R1-8B** | 2.24±1.20 | 2.80±1.41 | 2.88±1.67 | 2.64±1.33 |
| **Overall** | **2.99±1.44** | **3.61±1.25** | **3.54±1.48** | **3.38±1.25** |

Table S2 summarizes the mean and standard deviation of accuracy scores assigned independently by three expert physician reviewers across all answer generation models. Scores reflect each reviewer’s assessment of the factual accuracy of LLM-generated answers on a 5-point Likert scale (1 = completely inaccurate; 5 = completely accurate). Physician-assigned score refers to the mean of the three expert physician reviewers' ratings per response.

### Table S3. Mean and standard deviation of scores assigned by LLM-based evaluators under the reference-free scoring setting for each answer-generation model

| **Evaluation Methods** | **Answer Generation Models** | | | | | | **Statistical Test** |
| --- | --- | --- | --- | --- | --- | --- | --- |
|  | **Phi-4-14B** | **Mistral-7B** | **Qwen2-7B** | **Gemma2-7B** | **Llama3.2-3.2B** | **DeepSeek-R1-8B** | **Friedman p** |
| **Physician-Assigned Score** | 3.53±1.25 | 3.43±1.22 | 3.40±1.00 | 2.95±1.07 | 2.93±1.08 | 2.64±1.33 | 0.005 |
| **GPT-4 Evaluator** | 3.56±1.36 | 3.16±1.46 | 2.48±0.87 | 3.12±1.27 | 2.36±0.76 | 2.12±0.93 | p < 0.001 |
| **GPT-4o Evaluator** | 3.92±1.29 | 3.76±1.16 | 3.4±0.96 | 3.44±1.19 | 3.12±1.01 | 2.60±1.15 | p < 0.001 |
| **Gemma3-27B Evaluator** | 4.40±1.04 | 4.60±0.58 | 4.28±0.84 | 4.32±0.95 | 4.28±0.68 | 3.60±1.32 | 0.015 |
| **DeepSeek-R1-14B Evaluator** | 4.28±0.68 | 4.56±0.58 | 4.20±0.58 | 4.28±0.68 | 4.08±0.70 | 3.68±1.03 | 0.007 |
| **Qwen3-32B Evaluator** | 4.36±1.15 | 4.24±0.88 | 3.92±1.08 | 3.92±1.15 | 3.84±0.90 | 2.76±1.33 | p < 0.001 |
| **Llama3.3-70B Evaluator** | 4.48±1.12 | 4.84±0.37 | 4.76±0.44 | 4.56±0.87 | 4.48±0.51 | 3.76±1.39 | 0.002 |

### Table S4. Inference time and cost per response for LLM evaluators (Mean ± SD)

| **LLM evaluators** | **Inference Time (s/response)** | **Cost* (USD/response)** |
| --- | --- | --- |
| **GPT-4 Evaluator** | 1.68 ± 0.66 | (8.30 ± 1.40) × 10⁻³ |
| **GPT-4o Evaluator** | 1.51 ± 0.57 | (2.30 ± 0.37) × 10⁻³ |
| **Gemma3-27B Evaluator** | 13.08±4.95 | - |
| **DeepSeek-R1-14B Evaluator** | 21.59±3.9 | - |
| **Qwen3-32B Evaluator** | 30.6±8.96 | - |
| **LLaMA3.3-70B Evaluator** | 203.43±19.06 | - |

* Cost is approximated based on the number of tokens used per response.

### Table S5. Verbosity bias in LLM-based evaluation: Spearman correlation between evaluator scores and answer length under reference-guided scoring.

| **Correlation** | **Answer Length (words)** | |
| --- | --- | --- |
|  | Spearman ρ | p value |
| **Physician-Assigned Score** | 0.131 | 0.1 |
| **GPT-4 Evaluator** | 0.025 | 0.8 |
| **GPT-4o Evaluator** | 0.044 | 0.6 |
| **Gemma3-27B Evaluator** | 0.008 | 0.9 |
| **DeepSeek-R1-14B Evaluator** | 0.162 | 0.05 |
| **Qwen3-32B Evaluator** | 0.079 | 0.3 |
| **LLaMA3.3-70B Evaluator** | 0.054 | 0.5 |

As shown in Table S5, the low correlation between evaluator scores and answer length suggests that verbosity bias is not a major concern.
